# Covid-19 related deaths among doctors in India

**DOI:** 10.1101/2020.09.28.20202796

**Authors:** Aanandita Kapoor, Krishan Mohan Kapoor

## Abstract

**Background:** India has the most number of COVID-19 cases in the world currently, second only to the USA. The COVID-19 pandemic has caused high mortality not only in patients but also health care providers. In this paper, our aim is to analyze the cases of deaths in Indian doctors due to COVID-19 infection.

**Methods:** The details of data were taken from the list of the deceased doctors in India due to COVID-19 infection, which was compiled by the Indian Medical Association (IMA), the top body of Indian doctors practicing modern allopathic medicine. The key data fields of age, specialty, and geographical location of the deceased doctors were extracted from the given list, and analysis was performed.

**Results:** A total of 382 COVID-related deaths and 2174 infections were reported amongst doctors in India till 10 September 2020, with a case fatality rate of 16.7% among Indian doctors, which was ten times the CFR of 1.7% in the general population. Among the practicing doctors, after excluding the resident doctors and house surgeons, the CFR was 36.4%, which is almost 22 times more than what was seen in general population of India. The average age of COVID-related deaths in Indian doctors was 60.8 years, with a median age of 60; 62% of deaths among doctors were in the above 60 years, age group. The maximum number of deceased doctors were amongst general practitioners 225(58.9%). Among the specialists, most deaths were seen in paediatricians 26(6.8%), medical specialists 24(6.3%), general surgeons 22(5.8%), obstetricians & gynecologists 16(4.2%), and anesthesiologists 14(3.7%). The highest COVID-19 related deaths in doctors were seen in the Indian states of Tamil Nadu, Karnataka, Andhra Pradesh, Gujarat and Maharashtra in that order.

**Conclusions:** The mortality rate is very high among doctors in India compared to the general population. The average age of COVID-19 related death was 60 years among doctors. General practitioners and 60 years+ doctors are at a much higher risk of mortality among the doctors. The states with the high number of COVID-19 cases in India, also had a higher number of doctor deaths.

## Introduction

The COVID-19 pandemic has infected more than 4.5 million people in India, with an overall case fatality rate of 1.7% as of 10 September 2020. Doctors are at the frontline of healthcare delivery during the COVID-19 pandemic and are likely to get exposed to the infection^1^. The doctors treating and interacting with patients during these times are at a very high risk of contracting the infection, leading to a possible higher mortality rate than the general public. Doctors and other healthcare workers continue to selflessly face the risk of infection to help patients and colleagues^2^. COVID-19 related deaths among doctors have been reported from all over the world, including India. The characteristics of physician deaths in India from COVID infection were investigated in this study.

## Materials and methods

The data was collected from the list of deceased doctors released by the Indian Medical Association, IMA, the top body of Indian doctors practicing modern allopathic medicine. The list included names, age, specialization of the deceased doctors, and the state where they belonged. This list was presented to the Indian Government by IMA to give information about Indian doctors’ deaths. The list was updated last till 10 September. Corresponding data for the Indian population till 10 September was taken from a website www.worldometer.com. The mean and median of the age were calculated and the number of cases in each age group of doctors. The number of deceased doctors was also calculated for each medical specialty. In the list where the job designation was mentioned in doctors’ specialty, such doctors were grouped in the general practitioner category. The state-wise mortality figures were also calculated from the list. No ethics board approval was needed for this research paper, as this information was in the public domain.

## Results

Total COVID-related deaths in the Indian population till 10 September was 76,304 out of 4,559,725 total Covid-19 positive cases with a case fatality rate CFR of 1.7%. In all, 382 COVID related deaths were reported amongst doctors in India. These cases were out of 2174 total infected cases reported among doctors till 10 September 2020, a CFR of 16.7% among Indian doctors.

The Indian doctors infected with COVID-19 formed a very small percentage of the total cases in the population (2174 out of 76304 cases; 0.05%), but while calculating the deaths, this percentage increased sharply compared to deaths among the general population (382 out of 76304; 0.5%). The case fatality rate among Indian doctors at 16.7% was nearly ten times more than the general population (CFR 1.7%) on 10 September 2020.

Out of 2174 doctors who got infected with COVID 19, 1023(47%) were practicing doctors while 827(38%) were the resident doctors, and 324(15%) were house surgeons. The total deaths among resident doctors/ house surgeons were 10 out of 1151 infections, giving a CFR of 0.9%. On the other hand, deaths among practicing doctors were 372 out of 1023 infections, with a CFR of 36.4%, which is almost 22 times higher than the CFR of 1.7% in India’s general population.

The age range of the deceased doctors was 24-88 years (Figure 1). The percentage of deaths below 30 was 1%, below 40 years was 5.8%, below 50 years it was 11.8%. The percentage of deaths below 60 was 38%, while 62% of deaths among doctors were above 60. The average age of doctors at death was 60.8 years, with a median age of 60 years.

**Figure 1:**
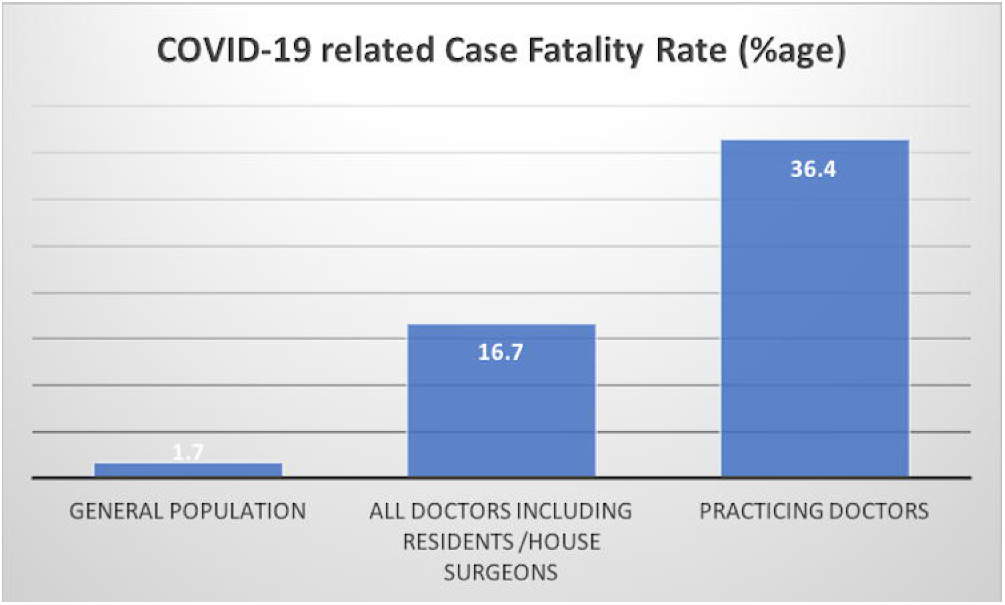
Case fatality rate was found to be very high among practicing doctors in India.

The majority of the deceased, 225(58.9%), were general practitioners (Figure 2). Among the specialists, the maximum brunt of COVID-19 was borne by paediatricians 26(6.8%), medical specialists 24(6.3%), general surgeons 22(5.8%), obstetricians & gynecologists 16(4.2%), and anaesthesiologists 14(3.7%). Amongst other specialists who lost their lives, orthopaedic surgeons 9(2.4%), ENT surgeons 8(2.1%), radiologists 5(1.3%), ophthalmologists 5(1.3%), and psychiatrists 4(1%) were major groups. There were three deaths (0.8%), each amongst pathologists, neurologists, and dentists. The super specialists who lost their lives comprised of neurologists 3(0.8%), cardiac surgeons 2(0.5%), urologists 2(0.5%), oncologist 1(0.26%), neurosurgeon 1(0.26%) and cardiologist 1(0.26%). The state-wise distribution of doctor deaths shows the highest deaths in Tamil Nadu, Andhra Pradesh, Gujarat, Maharashtra, and Karnataka (Figure 3). Tamil Nadu had the highest number of 61 deaths among the states.

**Figure 2:**
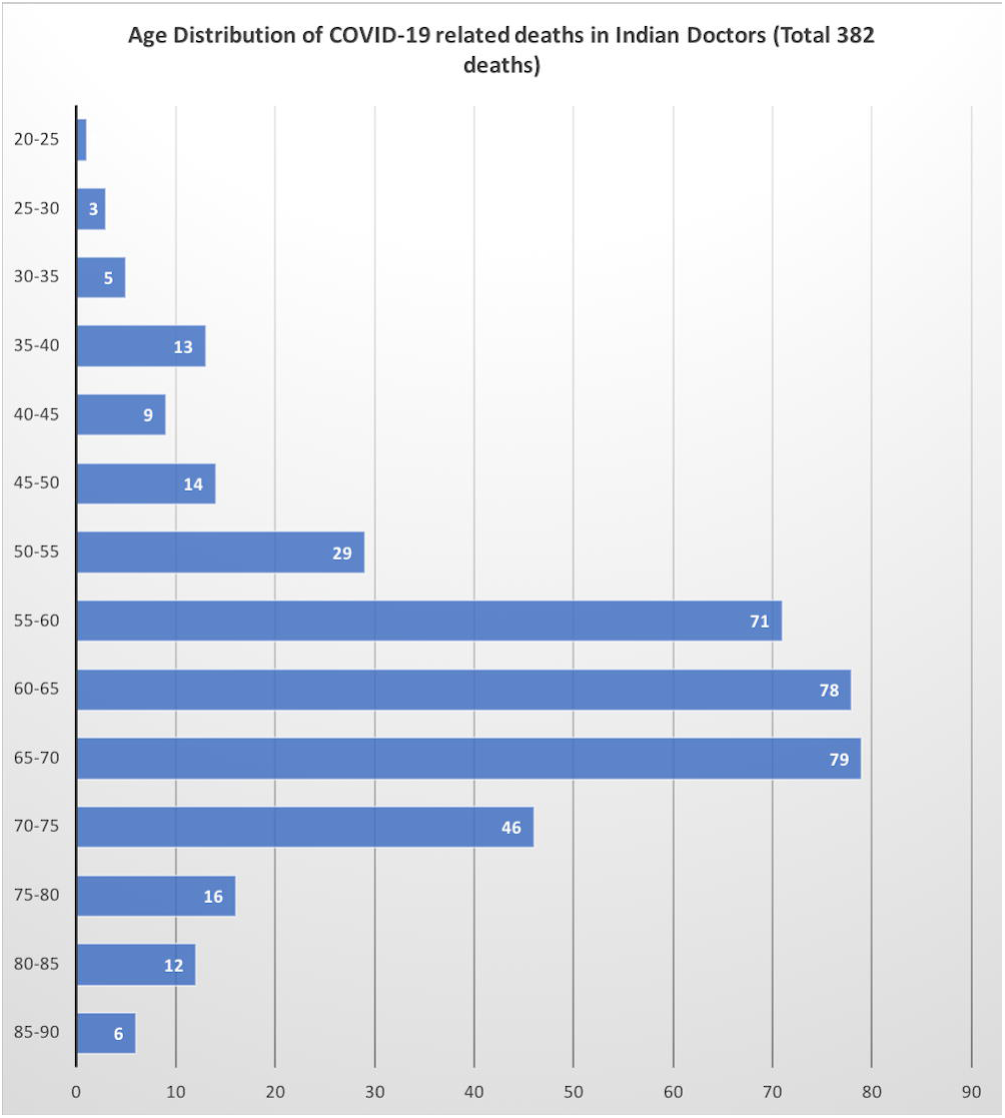
Age-specific deaths showing high mortality figures in 60 years and above age among Indian doctors.

**Figure 3:**
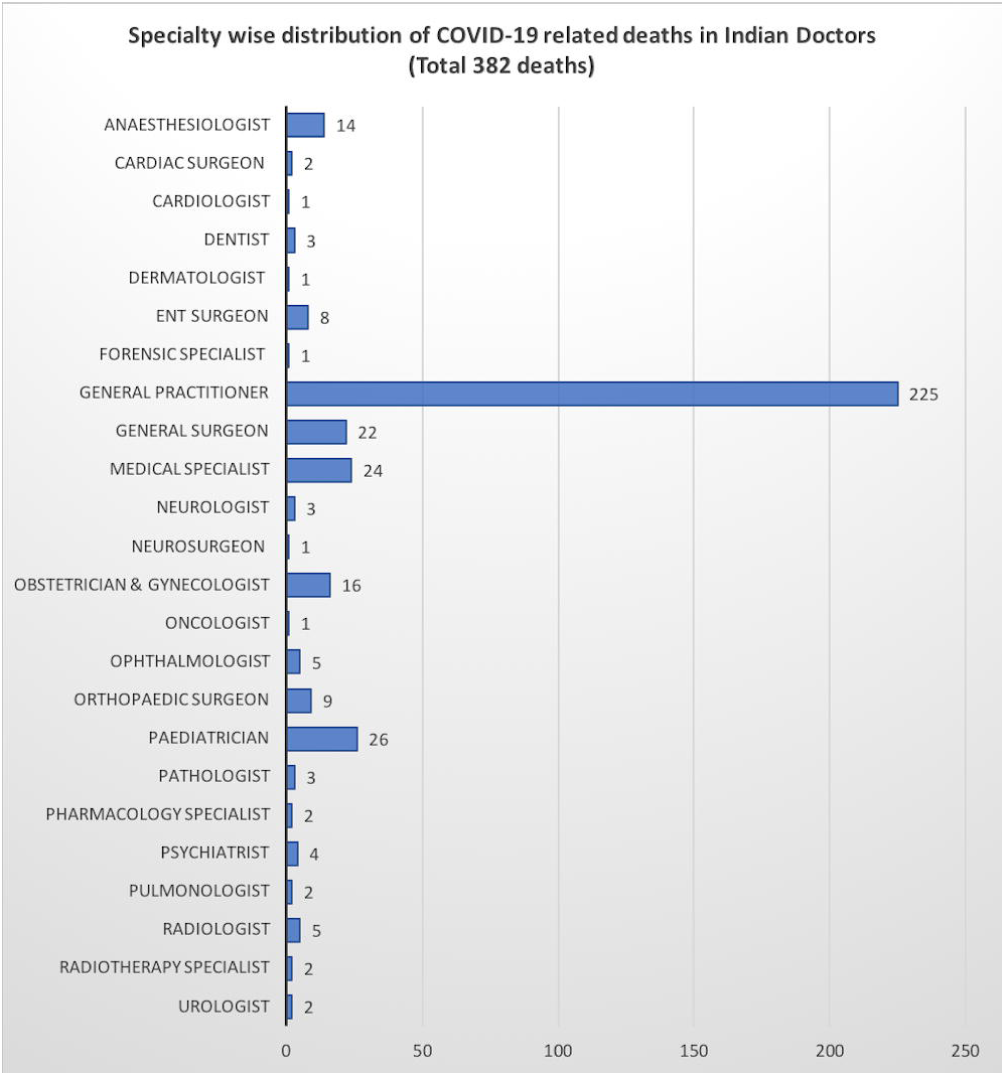
Specialty specific deaths showing high mortality figures among general practitioners, paediatricians and medical specialists.

**Figure 4:**
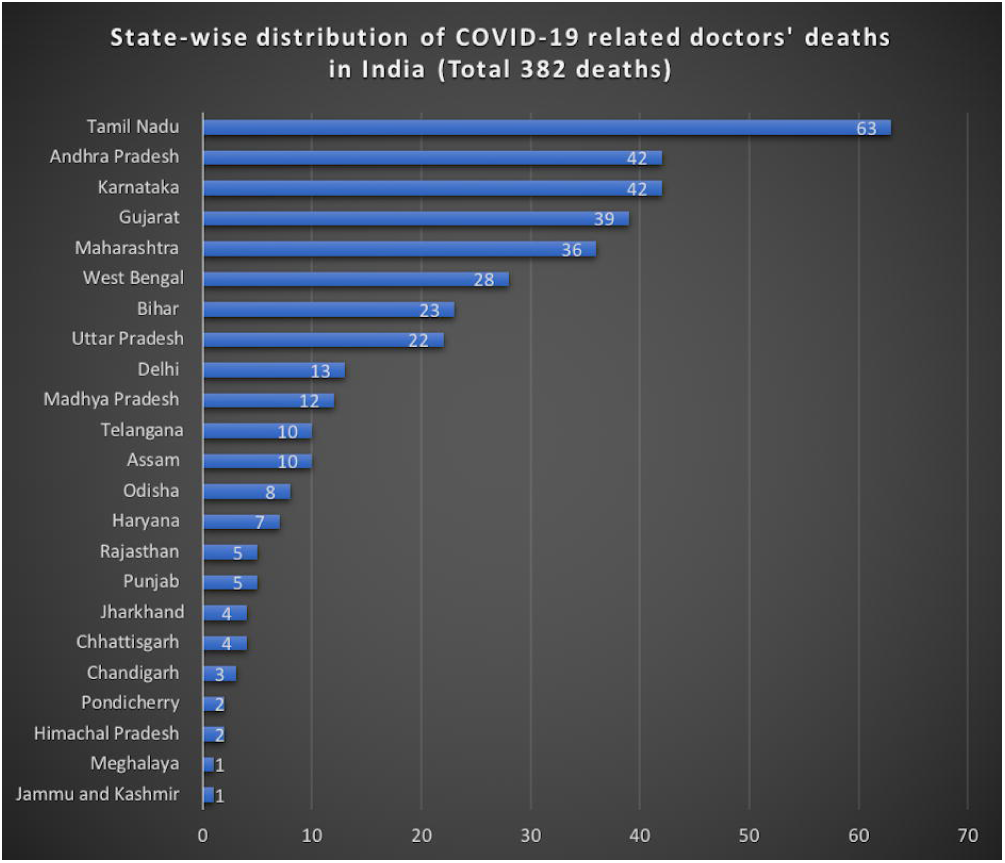
State-wise distribution of deaths showing higher mortality among doctors in states with high COVID-19 patients.

## Discussion

Physician deaths from COVID-19 vary between different countries due to disease outbreak timing, variation in public health resources, governmental policies and controls, enforcement of quarantine, social distancing and mask wear, and the rate of testing being performed, available medical supplies, technology & PPEs^3^. In an earlier study reporting 198 physician deaths, countries with the most reported deaths among doctors were: Italy (79/198; 40%), Iran (43/198; 22%), China (16/198; 8%), Philippines (14/198; 7%), United States (9/198; 5%), Indonesia (7/198; 4%) and France (6/198; 3%)^3^. An estimated 3000 healthcare workers were infected in China, and at least 22 had died^4 5^. In the present study, 382 confirmed deaths among Indian doctors were analyzed from the data made available by the Indian Medical Association. Indians doctors have a higher mortality risk due to COVID-19 infection as Indian healthcare workers were found to have more than three times mortality in the UK NHS than their Caucasian counterparts^6^.

In our study, the majority of doctors’ deaths were in the general practitioners’ group. In India, healthcare workers, especially in general practice, are at higher risk of getting COVID-19 infection due to the relatively large volume of patients attended and frequently congested working conditions at workplaces^1^. The higher number of doctors’ deaths in the states having more COVID-19 infections also indicate this trend. In another study, emergency medicine and general practitioners accounted for approximately 40% of deaths^7^. Physicians working in the airway such as dentists, ENT surgeons, and anaesthesiologists are also at high risk for COVID-19 infection, and this group comprised 25/ 382 (6.5%) of the physician deaths in India. Three ophthalmologists in the Central Hospital of Wuhan, Hubei, lost their lives due to COVID-19 as it has been found that SARS-CoV-2 can be detected in tears and conjunctival secretions of patients^8^. Studies have suggested that with no eye protection, SARS-CoV-2 could also possibly be spread by aerosol contact with conjunctiva and cause infection of doctors^9^. In our study, five ophthalmologists have lost their lives in India till now.

Certain factors attributed to death in frontline doctors include lack of adequate PPE, an inadequate technique of donning and doffing of PPE, and non-disclosure by patients of their exposure to possible COVID-19 infection, excessive working hours of doctors, and poor doctor-patient ratio^10^. Individual comorbidities in doctors, including advanced age, diabetes mellitus, cardiovascular diseases, chronic lung disease, or immunocompromised states, contribute to the higher death rates^11^. In our study, 62% of deaths were seen in doctors aged 60 years or above. The higher proportion of deaths in NHS, UK, that has occurred in doctors aged >60 indicates that caution should be used about placing these doctors in frontline clinical roles due to a higher risk of contracting COVID-19 infection^12^.

The other recognized risk factors among doctors are close contact in physical examination and therapy of infected patients, direct contact with body fluid and excreta from patients, suctioning of airways, endotracheal intubation, and cardiopulmonary resuscitation. However, the doctors’ most dangerous situation is when they face the super-spreader, whose transmission ability is just overwhelming^13^.

In the case of infections like COVID-19, the precautionary principle should be used for frontline health workers, and a properly fitted respirator or masks should be used. There should be a uniform policy regarding the use of personal protective equipment where health workers’ occupational health and safety is a high priority^14^. Telemedicine reduces risk to the treating physicians and reinforces the health systems with the health care workers quarantined at home after exposure to COVID-19 infection^15^.

Liberating clinicians from other tasks and commitments allow them to focus on their immediate needs. Providing food, rest breaks, decompression time, and adequate time off from work may be as important as the protocols and protective equipment during a protracted battle against COVID-19 infection^4^. Many doctors’ bodies and associations have given their specialties guidelines to safeguard the member physicians against COVID-19 infection^16 17^. The British Medical Association BMA has written to the UK chancellor of the exchequer to request that all NHS workers receive full ‘death in service’ cover after an announcement by the Scottish government of a comprehensive ‘death in service’ package for all its NHS workers^18^. Similar support by the Indian government to doctors in India will go a long way in motivating them to give their best.

During this pandemic, the circumstances have shown that robust healthcare systems are made of more than bricks and mortar, data flow or supply chains, and highly skilled healthcare workers are essential members of healthcare systems. The general public recognizes this fact during the celebration of healthcare workers on social media around the world^19^. Protecting and empowering all healthcare workers will be very important for reducing the impact of this pandemic and building deeper resilience into healthcare systems^20^. Whether the government prevention strategy aims to suppress or mitigate, building healthcare capacity should be a top priority that also includes protecting healthcare workers as our most valuable resource^21^. A physically and mentally healthy and well-equipped healthcare workforce is essential for a country to effectively manage COVID-19 cases. Lessons can be learned from past epidemics to introduce special working arrangements to help protect healthcare workers from the infections^22^.

## Conclusions

In India, physicians from almost all medical and surgical specialties have succumbed to COVID-19 infection. With more cases happening with every passing day, the number of physician fatalities is likely to increase in the coming months. Doctors who were 60 years of age or older accounted for 62% of COVID-19-related deaths among doctors. General practitioners accounted for nearly 59% of COVID-19 related deaths among doctors in India.

## Limitations

Though the number of deaths may be more accurate to ascertain, the number of physician infections from COVID-19 could be under-reported given the fast-changing course of the pandemic, and because all the doctors are not tested. We could not discern if the physicians died from COVID-19 were managing the COVID-19 cases or not. Pre-existing medical morbidities were also not reported on the list. The paper also does not mention infections and deaths in the nursing and allied health staff due to a lack of data.

## Data Availability

The data used in the paper is taken from list of the deceased doctors made by Indian Medical Association.

https://www.ima-india.org/ima/

## Abbreviations

CFR: Case Fatality Rate
PPE: Personal Protective Equipment
IMA: Indian Medical Association
BMA: British Medical Association

## Competing interests

None

## Funding

None

## Notes

### Competing Interest Statement

The authors have declared no competing interest.

### Funding Statement

No funding received.

## Bibliography

1. Jayadevan R. A Hundred Lives Lostl J: Doctor Deaths in India During the Times of COVID-19. 2020;(July). doi:10.20944/preprints202007.0346.v1

2. Harkin DW. Covid-19: Balancing personal risk and professional duty. BMJ. 2020;369(April):2020. doi:10.1136/bmj.m1606

3. Ing EB, Xu QA, Salimi A, Torun N. Physician deaths from corona virus (COVID-19) disease. Occup Med (Lond). 2020;70(5):370–374. doi:10.1093/occmed/kqaa088

4. Adams JG, Walls RM. Supporting the Health Care Workforce during the COVID-19 Global Epidemic. JAMA - J Am Med Assoc. 2020;323(15):1439–1440. doi:10.1001/jama.2020.3972

5. Chen W, Huang Y. To Protect Healthcare Workers Better, To Save More Lives. Anesth Analg. March 2020:10.1213/ANE.0000000000004834. doi:10.1213/ANE.0000000000004834

6. Jankowski J, davies A, English P, et al. Risk Stratification tool for Healthcare workers during the CoViD-19 Pandemic; using published data on demographics, co-morbid disease and clinical domain in order to assign biological risk. medRxiv. January 2020:2020.05.05.20091967. doi:10.1101/2020.05.05.20091967

7. Kursumovic E, Lennane S, Cook TM. Deaths in healthcare workers due to COVID-19: the need for robust data and analysis. Anaesthesia. 2020;75(8):989–992. doi:10.1111/anae.15116

8. Xia J, Tong J, Liu M, Shen Y, Guo D. Evaluation of coronavirus in tears and conjunctival secretions of patients with SARS-CoV-2 infection. J Med Virol. 2020;92(6):589–594. doi:10.1002/jmv.25725

9. Xiao J, Fang M, Chen Q, He B. SARS, MERS and COVID-19 among healthcare workers: A narrative review. J Infect Public Health. 2020;13(6):843–848. doi:10.1016/j.jiph.2020.05.019

10. Iyengar KP, Ish P, Upadhyaya GK, Malhotra N, Vaishya R, Jain VK. COVID-19 and mortality in doctors. Diabetes Metab Syndr Clin Res Rev. 2020;14(6):1743–1746. doi:https://doi.org/10.1016/j.dsx.2020.09.003

11. Jordan RE, Adab P, Cheng KK. Covid-19: risk factors for severe disease and death. BMJ. 2020;368. doi:10.1136/bmj.m1198

12. Majeed A, Molokhia M, Pankhania B, Asanati K. Protecting the health of doctors during the COVID-19 pandemic. Br J Gen Pract. 2020;70(695):268 LP - 269. doi:10.3399/bjgp20X709925

13. Ebrahim SH, Memish ZA. COVID-19: preparing for super-spreader potential among Umrah pilgrims to Saudi Arabia. Lancet (London, England). 2020;395(10227):e48–e48. doi:10.1016/S0140-6736(20)30466-9

14. Chughtai AA, Seale H, Islam MS, Owais M, Macintyre CR. Policies on the use of respiratory protection for hospital health workers to protect from coronavirus disease (COVID-19). Int J Nurs Stud. 2020;105:103567. doi:https://doi.org/10.1016/j.ijnurstu.2020.103567

15. Moazzami B, Razavi-Khorasani N, Dooghaie Moghadam A, Farokhi E, Rezaei N. COVID-19 and telemedicine: Immediate action required for maintaining healthcare providers well-being. J Clin Virol. 2020;126:104345. doi:https://doi.org/10.1016/j.jcv.2020.104345

16. Huang Z, Zhao S, Li Z, et al. The Battle Against Coronavirus Disease 2019 (COVID-19): Emergency Management and Infection Control in a Radiology Department. J Am Coll Radiol. 2020. doi:https://doi.org/10.1016/j.jacr.2020.03.011

17. Kapoor KM, Chatrath V, Boxley SG, et al. COVID-19 Pandemic: Consensus Guidelines for Preferred Practices in an Aesthetic Clinic. Dermatol Ther. 2020;n/a(n/a):e13597. doi:10.1111/dth.13597

18. Rimmer A. Covid-19: BMA demands full death in service benefits for UK doctors. BMJ. 2020;369. doi:10.1136/bmj.m1634

19. Koh D. Occupational risks for COVID-19 infection. Occup Med (Chic Ill). 2020;70(1):3–5. doi:10.1093/occmed/kqaa036

20. Smith C. The structural vulnerability of healthcare workers during COVID-19: Observations on the social context of risk and the equitable distribution of resources. Soc Sci Med. 2020;258(June):113119. doi:10.1016/j.socscimed.2020.113119

21. Godderis L, Boone A, Bakusic J. COVID-19: a new work-related disease threatening healthcare workers. Occup Med (Lond). 2020;70(5):315–316. doi:10.1093/occmed/kqaa056

22. Sim MR. The COVID-19 pandemic: Major risks to healthcare and other workers on the front line. Occup Environ Med. 2020;77(5):281–282. doi:10.1136/oemed-2020-106567

